# Markers of Inflammation are Associated with Symptoms and Neighborhood Deprivation in Black Adults with Heart Failure

**DOI:** 10.1101/2025.05.10.25327361

**Authors:** Brittany Butts, Christopher Herring, Andrea Madariaga, Lucinda J Graven, Melinda K Higgins, Sandra B Dunbar

## Abstract

**Background:** Black adults face a higher prevalence of heart failure (HF), with greater symptom burden and earlier onset compared to other populations. Systemic inflammation and socioeconomic factors, including neighborhood deprivation, contribute to these disparities. Understanding the interplay between inflammation, HF symptoms, and social determinants of health is critical for addressing inequities in HF outcomes.

**Objectives:** This study examines associations between inflammatory biomarkers and physical and psychological symptoms in Black adults with HF and explores the impact of neighborhood deprivation on these factors.

**Methods:** Black adults with HF (N=41) were enrolled in this cross-sectional study. Blood samples were collected using Mitra Microsampling for biomarker analysis, including cytokines, chemokines, and xanthine oxidase (XO) activity. Symptoms were assessed using validated measures for dyspnea, fatigue, anxiety, depression, stress, and sleep disturbance. Neighborhood deprivation was evaluated using the Area Deprivation Index.

**Results:** Elevated XO activity was significantly associated with dyspnea severity (β = 0.75, p < .001). Chemokines linked to T cell activation (e.g., C-C motif ligand[CCL]-11, CXC motif ligand[CXCL]-8) correlated with HF symptoms and psychological distress, including anxiety and perceived stress. Higher neighborhood deprivation scores were associated with increased stress, sleep disturbance, and inflammatory biomarkers (e.g., interleukin[IL]-4, vascular cell adhesion molecule[VCAM]-1).

**Conclusions:** This study highlights the role of inflammation and neighborhood deprivation in HF symptomatology among Black adults. Targeting oxidative stress and inflammatory pathways, alongside addressing social determinants of health, may reduce symptom burden and improve outcomes in this population.

## Introduction

There is a higher prevalence of heart failure (HF) among Black individuals compared to other racial groups, ^1^ and HF manifests earlier in life among Black individuals compared to White counterparts, leading to a higher burden of disease at a younger age. ^1^ Further, Black adults with HF experience higher mortality and readmission rates compared to other racial groups. ^1,2^ Health inequities in Black adults with HF often stem from structural racism and social determinants of health, such as the lived environment, access to care, and socioeconomic status, contributing to chronic stress, poor sleep, and systemic inflammation. ^3^ This chronic inflammation exacerbates cardiovascular issues, contributing to the disproportionate burden of HF in this population. ^4^ Despite disparities in HF severity and morbidity among Black adults as compared to White adults, relationships between inflammation and symptom burden are not well characterized.

HF is a complex syndrome involving the interplay of pathophysiologic processes, such as inflammation and myocardial remodeling. Worsening HF has been associated with overactivation of inflammatory pathways, including oxidative stress, ^5^ and dysregulation of the adaptive immune system, including T cell activation. ^6^ T cell activation contributes to HF progression through cytokine release leading to cardiac fibrosis and hypertrophy. ^7^

Elevated oxidative stress, which is associated with increased chronic stress, has been shown to play a key role in the development and progression of HF by inducing apoptosis, hypertrophy, and inflammation in myocardial tissue which leads to impaired ventricular function. ^8, 9^ Xanthine oxidase (XO) activity, an enzyme that catalyzes the conversion of xanthine to uric acid and serves as an indicator of oxidative stress, was found to be higher in Black individuals with resistant hypertension compared to White individuals. ^10^ Increased levels of reactive oxygen species have been associated with dysregulated T-cell activation, signaling, and proliferation. ^11^ T-cell overactivation contributes to HF progression through excessive cytokine release, leading to cardiac fibrosis and hypertrophy.

HF is characterized by physical symptoms that have been shown to predict morbidity and mortality. Physical symptoms, especially fatigue, are often closely linked with depressive symptoms in persons with HF, and together can have a compounding effect leading to impaired self-care and worse HF outcomes. ^12^ Depressive symptoms contribute to HF outcomes, such as quality of life, HF-specific health status, cardiac events, mortality, rehospitalization, and utilization of health care resources. ^13^ However, the pathophysiologic mechanisms driving symptoms are not well understood.

There is a critical need to understand pathophysiologic mechanisms associated with symptoms and drivers of health disparities in Black adults with HF. Thus, the purpose of this pilot study is to examine the relationship between inflammatory biomarkers and symptoms in Black adults with HF and explore the role of neighborhood disadvantage on HF symptoms and inflammation.

## Materials and Methods

### Study Design and Sample

This cross-sectional observational study enrolled 41 self-identified Black adults living with HF, as previously described. ^14^ Participants were recruited from HF clinics at Emory University Hospitals from March 2021 to October 2022. The general inclusion criteria were: 1) Self-reported Black race including African American, Afro-Caribbean, and African; 2) between 30-80 years of age; 3) able to read, write, and understand English. The exclusion criteria for the study included: 1) diagnosed with an uncontrolled major mental disorder (i.e., schizophrenia, bipolar disorder, major depression) as noted in the electronic medical records; 2) severe chronic kidney disease (GFR <30).

This study was performed under research protocols approved by the Institutional Review Boards of Emory University. Informed consent was obtained remotely via secure electronic measures or via telephone, based on individual preference and electronic accessibility. Study data were collected and managed using REDCap (Research Electronic Data Capture). ^15,16^ Sociodemographic data were collected by self-report and clinical data were collected via the electronic medical record. Comorbidities were quantified using the Charlson Comorbidity Index (CCI), which estimates mortality risk by considering age, number of comorbidities, and severity of each comorbidity. ^17^ A higher CCI score is associated with a higher 1-year and 10-year mortality rate. ^17^

### Specimen Acquisition and Processing

Blood was self-collected by participants using dried blood spots via Mitra Microsampling after an overnight fast, as previously described. ^14,18^ Briefly, samples were extracted from the microsampling devices with a buffer (0.05% Tween® 20 [polyethylene glycol sorbitan monolaurate] in phosphate-buffered saline) with a protease inhibitor (cOmplete^™^, Mini Protease Inhibitor Cocktail, Roche, Basel, Switzerland) and stored at -80°C until analysis.

### Measures

#### Biomarkers

Inflammatory and cardiometabolic biomarkers were measured via multiplex immunoassays (Target 48 Cytokine and Target 96 Cardiometabolic panels, Olink®, Uppsala, Sweden). Biomarkers included interleukin (IL) family cytokines (IL-1β, IL-6, IL-15, IL-4, IL- 18), matrix metalloproteinases (MMP-1, MMP-12), C-C motif chemokine ligand (CCL) chemokines (CCL-11, CCL-19), C-X-C motif chemokine ligand (CXCL) chemokines (CXCL-3, CXCL-8, CXCL-11), vascular cell adhesion molecule (VCAM)-1, and C1q/tumor necrosis factor-related protein 1 (CTRP1). Samples were randomized on plates using a random number generator and were run in triplicate. A calibrator of pooled human plasma sample with spiked-in recombinant antigens for proteins with low endogenous levels was used to ensure all proteins were detected within the limit of quantification. Intensity normalization was performed with the following steps for each assay: 1. the overall median value for all samples and plates was calculated, 2. the plate-specific median value was calculated, 3. the plate-specific median was subtracted from every sample of the plate, which centralized the median to 0, and 4. for each assay, the overall median value was added to every sample in the project which equals centralizing to the overall median.

XO activity was determined by a coupled enzyme assay (Xanthine Oxidase Activity Assay Kit, Sigma-Aldrich^®^, St. Louis, MO), which resulted in a fluorometric product, proportional to the hydrogen peroxide generated by xanthine oxidase (B). Standard serial dilutions were used to create a standard curve and determine maximum activity values.

Absorbance at 535/587 nm was measured after 2 minutes of reaction time (time 1, T1) and again at 20 minutes (time 2, T2). The molar concentration of H2O2 generation (B) between T1 and T2 was calculated using the standard curve. XO activity was calculated as [(B) x sample dilution factor]/[(T2 – T1) x sample volume (mL)] and was reported as nmol/min/mL, where one milliunit of XO is defined as the amount of enzyme that catalyzes the oxidation of xanthine yielding 1.0 µmole of uric acid and hydrogen peroxide per minute at 25°C.

#### Symptom Questionnaires

The Patient-Reported Outcomes Measurement Information System (PROMIS) <u>Dyspnea</u> Severity SF (v1.0) was used to provide a standardized assessment of dyspnea. This 10-item questionnaire assessed the severity of shortness of breath an adult experienced in response to various physical activities (i.e., walking, dressing, making a bed, etc.) over the past seven days. Raw scores were converted into standardized T scores with a mean of 50 and a standard deviation of 10 (calibrated using a sample of COPD patients) using the HealthMeasures Scoring Service.^15^ Reliability and validity of this measure has been established.^16^ <u>HF Somatic Symptoms</u> were measured with The Heart Failure Somatic Perception Scale v3 (HFSPS), a four-factor, 18- item valid and reliable measure of HF symptom perception and burden in adults with symptomatic HF. ^19^ The HFSPS total, dyspnea, and early and subtle subscale scores are associated with physical limitations and predicted HF event-free survival. <u>Fatigue</u> was measured by the Multidimensional Fatigue Inventory (MFI-20), ^20^ a 20 item Likert scale instrument used successfully with chronically ill patients. The MFI-20 has good internal consistency and test-retest scores. ^20^ The state scale of the State-Trait <u>Anxiety</u> Inventory (STAI) ^21^ was used to assess how participants feel at the moment and is a sensitive indicator of transitory anxiety versus anxiety proneness. ^21^ Higher scores indicate higher levels of anxiety; STAI scores ≥ 40, indicate clinically significant anxiety. ^21^ <u>Depressive Symptoms</u> were measured by the Center for Epidemiological Studies Depression Scale (CES-D). ^22^ The CES-D assesses various symptoms of depression including negative mood, decreased positive affect, and loneliness over the past seven days, and has shown to be a valid and reliable instruments, including with ethnically diverse samples. ^23^ The <u>Perceived Stress</u> Scale (PSS) is a widely used self-report questionnaire designed to assess an individual’s perception of stress in their life over the past month. ^24^ The scale consists of 10 items that measure the degree to which situations in one’s life are appraised as stressful. The items are based on feelings and thoughts related to stress, asking respondents to rate their frequency of experiences on a 5-point Likert scale, ranging from 0 (Never) to 4 (Very Often). The Perceived Stress Scale demonstrates robust construct validity through consistent correlations with related measures and exhibits strong internal consistency and test-retest reliability across various populations and settings. ^25^

#### Sleep Disturbance

The PROMIS Sleep Disturbance Scale is a validated self-reported questionnaire that assesses various aspects of sleep disturbance in individuals. ^26^ Comprising eight items, the scale covers different facets of sleep, including quality, depth, and regularity, with responses rated on a 5-point Likert scale. The scale has demonstrated strong validity and reliability across diverse populations. ^26^

#### Area Deprivation Index

Neighborhood socioeconomic disadvantage was measured by the 2021 Area Deprivation Index (ADI). ^27,28^ The ADI includes factors for theoretical domains of income, education, employment, and housing quality, and is identified by an individual’s nine-digit zip code.

Drawing from diverse census-based indicators encompassing income, education, employment, housing quality, and access to resources, the ADI integrates this information into a singular score for each area. ADI values are provided in percentile rankings at the block group level ranging from 1 to 100 for national scores and from 1 to 10 for state scores, with one as the lowest score and 100/10 as the highest level of deprivation.

### Data Analysis

All data were reviewed for data entry errors, normality assumptions, potential outliers, and missing data. Biomarker variables that did not meet normality assumptions were transformed using natural log (LN) logarithmic transformation. For continuous variables, descriptive data were presented as mean ± standard deviation or median (interquartile range), based on normality assumptions. Associations between study variables and covariates were analyzed using Pearson correlations for continuous variables, Student’s t-test for binomial variables, and one-way ANVOA for categorical variables. Effect size was calculated using Cohen’s d for T-tests and η^2^ for ANOVA. Linear regression analyses were performed to examine linear relationships between variables, controlling for covariates (including age, gender, creatinine levels, left ventricular ejection fraction, and comorbidities) based on hypothesized relationships within the study. All analyses were performed using SPSS version 28.

## Results

Participants (N=41) were 57 years of age (range 34-80) and 66% female (**Table 1**). The mean left ventricular ejection fraction (LVEF) was 33%. The majority of participants (n=26, 63%) had HF with reduced EF (LVEF<40%), 6 (15%) participants had HF with preserved EF (LVEF≥50%), and 9 (22%) had HF with mid-range EF (LVEF 40-49%). The most common comorbidities were hypertension, diabetes, stroke, and myocardial infarction (**Table 1**). Mean creatinine levels for this population were 1.16 ± 0.4 uM/ml, with a range of 0.6 to 2.15 uM/ml.

**Table 1.**
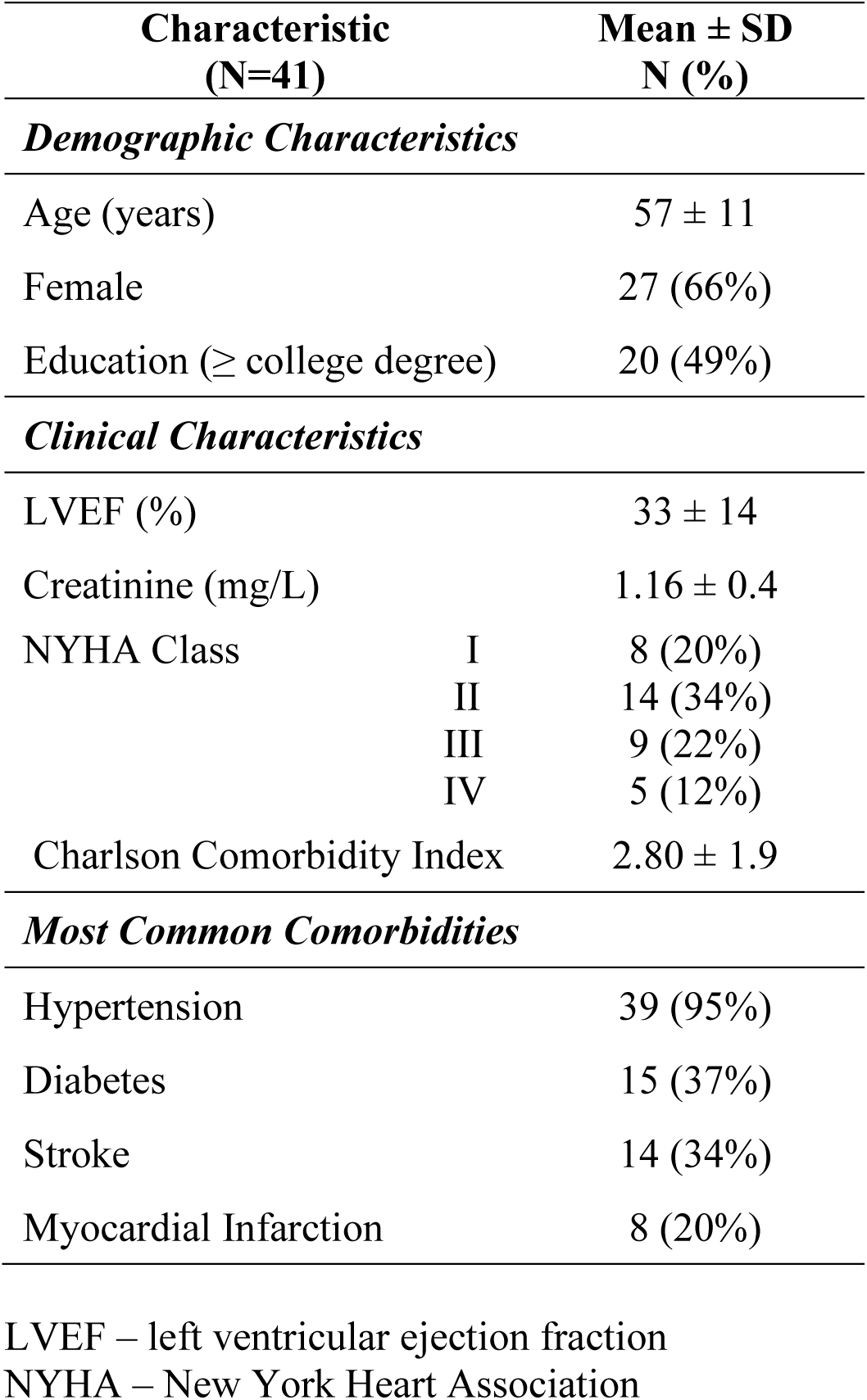
Demographic and Clinical Data.

| **Characteristic**<br>**(N=41)** |  | **Mean ± SD**<br>**N (%)** |
| --- | --- | --- |
| ***Demographic Characteristics*** |  |  |
| Age (years) | | 57 ± 11 |
| Female |  | 27 (66%) |
| Education (≥ college degree) | | 20 (49%) |
| ***Clinical Characteristics*** |  |  |
| LVEF (%) | | 33 ± 14 |
| Creatinine (mg/L) | | 1.16 ± 0.4 |
| NYHA Class | I | 8 (20%) |
|  | II | 14 (34%) |
|  | III | 9 (22%) |
|  | IV | 5 (12%) |
| Charlson Comorbidity Index | | 2.80 ± 1.9 |
| ***Most Common Comorbidities*** |  |  |
| Hypertension |  | 39 (95%) |
| Diabetes |  | 15 (37%) |
| Stroke |  | 14 (34%) |
| Myocardial Infarction |  | 8 (20%) |

LVEF – left ventricular ejection fraction
NYHA – New York Heart Association

Summary data for symptom questionnaires and biomarkers are presented in **Table 2** and **Table 3**, respectively. New York Heart Association (NYHA) Class IV was significantly higher than Class I for XO activity (8.43 ± 4.6 vs 0.71 ± 0.5 nmol/min/mL, η^2^=.34) and IL-6 (0.50 ± 0.1 vs 0.19 ± 0.1 pg/ml, η^2^=.34; **Supplementary Table 1**), with large effect sizes. LVEF was negatively associated with XO activity (r=-.362, p=.038).

**Table 2.** Symptom Questionnaires and Subscales.

| **Symptom** | **Mean ± SD** |
| --- | --- |
| ***Heart Failure Somatic Perception Scale (HFSPS)*** |  |
| HFSPS Total Score | 18.12 ± 15.2 |
| HFSPS Dyspnea Subscale | 5.08 ± 6.7 |
| HFSPS Subtle Early Subscale | 8.94 ± 6.0 |
| HFSPS Chest Discomfort Subscale | 1.81 ± 2.2 |
| HFSPS Edema Subscale | 2.25 ± 2.4 |
| ***Multidimensional Fatigue Inventory (MFI)*** |  |
| MFI Total | 50.09 ± 16.8 |
| MFI General Fatigue | 12.00 ± 3.7 |
| MFI Physical Fatigue | 12.26 ± 4.3 |
| MFI Reduced Activity | 9.94 ± 4.2 |
| MFI Reduced Motivation | 8.82 ± 4.0 |
| MFI Mental Fatigue | 7.51 ± 3.3 |
| ***PROMIS Measures*** |  |
| PROMIS Dyspnea Severity | 49.13 ± 12.2 |
| PROMIS Sleep Disturbance | 47.76 ± 12.4 |
| ***Psychological Symptoms*** |  |
| Perceived Stress Scale | 17.38 ± 10.3 |
| State-Trait Anxiety Index | 31.77 ± 14.1 |
| CES-Depression | 10.27 ± 11.7 |

CES – Center for Epidemiologic Studies
PROMIS - Patient-Reported Outcomes Measurement Information System

**Table 3.** Inflammatory and Cardiometabolic Biomarkers.

| **Biomarker** | **Mean ± SD**<br>**Median (IQR)** |
| --- | --- |
| Xanthine oxidase activity (nmol/min/mL) | 1.07 (1.26) |
| Matrix metalloproteinase-1 (pg/ml) | 541.40 (581.33) |
| Matrix metalloproteinase-12 (pg/ml) | 13.66 ± 9.8 |
| Interleukin-1β (pg/ml) | 25.43 (29.79) |
| Interleukin-4 (pg/ml) | 0.19 (0.16) |
| Interleukin-6 (pg/ml) | 0.35 (0.56) |
| Interleukin-15 (pg/ml) | 6.68 (4.10) |
| Interleukin-18 (pg/ml) | 573.66 (423.50) |
| C-C motif chemokine ligand-11 (pg/ml) | 1.97 (1.43) |
| C-C motif chemokine ligand-19 (pg/ml) | 3.74 (3.65) |
| C-X-C motif chemokine ligand-3 (pg/ml) | 1.52 (1.16) |
| C-X-C motif chemokine ligand-8 (pg/ml) | 10.41 (13.46) |
| C-X-C motif chemokine ligand-11 (pg/ml) | 1.18 (1.40) |
| Vascular cell adhesion molecule-1 (pg/ml) | 4.12 ± 0.6 |
| C1q/tumor necrosis factor-related protein 1 (pg/ml) | 4.10 ± 0.7 |

Physical symptoms differed by NYHA class, with lower NYHA class associated with lower symptoms (**Supplementary Table 1**). Age was negatively associated with CES-D (r=- .377, p=.034) and PROMIS Sleep Disturbance (r=-.445, p=.010), suggesting older age is associated with higher depressive symptoms and more sleep disturbance in this population.

### Area Deprivation Index

Mean National and State ADI scores were 61 ± 21 and 6 ± 2, respectively, indicating higher than average neighborhood deprivation levels. Associations between National and State ADI scores and outcome variables (symptoms, biomarkers) are presented as unadjusted models and models adjusting for age, sex, and comorbidities (**Table 4**). National and State ADI scores were positively associated with both PSS and PROMIS Sleep Disturbance scores. Additionally, National and State ADI scores were positively associated with inflammatory cytokines IL-4 and IL-18 and cardiometabolic biomarkers VCAM-1 and CTRP1.

**Table 4.** Adjusted and Unadjusted Effect Size Estimates for Area Deprivation Index and Associated Outcomes.

|  | Unadjusted<sup>1</sup> |  | Adjusted<sup>2</sup> |  |
| --- | --- | --- | --- | --- |
| | $\beta$ | p-value | $\beta$ | p-value |
| ***Perceived Stress Scale*** |  |  |  |  |
| ADI National | .538 | .007 | .659 | .003 |
| ADI State | .537 | .007 | .587 | .007 |
| ***PROMIS Sleep Disturbance*** |  |  |  |  |
| ADI National | .473 | .020 | .433 | .052 |
| ADI State | .481 | .017 | .434 | .039 |
| ***Interleukin-4 (pg/ml)*** |  |  |  |  |
| ADI National | .537 | .004 | .578 | .003 |
| ADI State | .588 | .001 | .582 | .002 |
| ***Interleukin-18 (pg/ml)*** |  |  |  |  |
| ADI National | .465 | .013 | .424 | .029 |
| ADI State | .456 | .015 | .429 | .020 |
| ***Vascular cell adhesion molecule-1 (pg/ml)*** |  |  |  |  |
| ADI National | .481 | .010 | .670 | <.001 |
| ADI State | .553 | .002 | .666 | <.001 |
| ***Clq/tumor necrosis factor-related protein 1 (pg/ml)*** |  |  |  |  |
| ADI National | .409 | .031 | .489 | .017 |
| ADI State | .450 | .016 | .512 | .008 |

1. Linear regression model with no covariates
2. Linear regression model adjusted for age, sex, and Charlson Comorbidity Index

### Associations Between Symptoms and Biomarkers

Associations between symptoms and biomarkers are presented as unadjusted models and models adjusting for age, sex, and comorbidities (**Table 5**). CCL-11 was positively associated with STAI, and CXCL-11 was positively associated with CESD. XO activity was positively associated with dyspnea severity. Dyspnea severity was positively associated with MMP-1 and MMP-12, enzymes that promote tissue remodeling and breakdown of collagen and elastin.

**Table 5.**
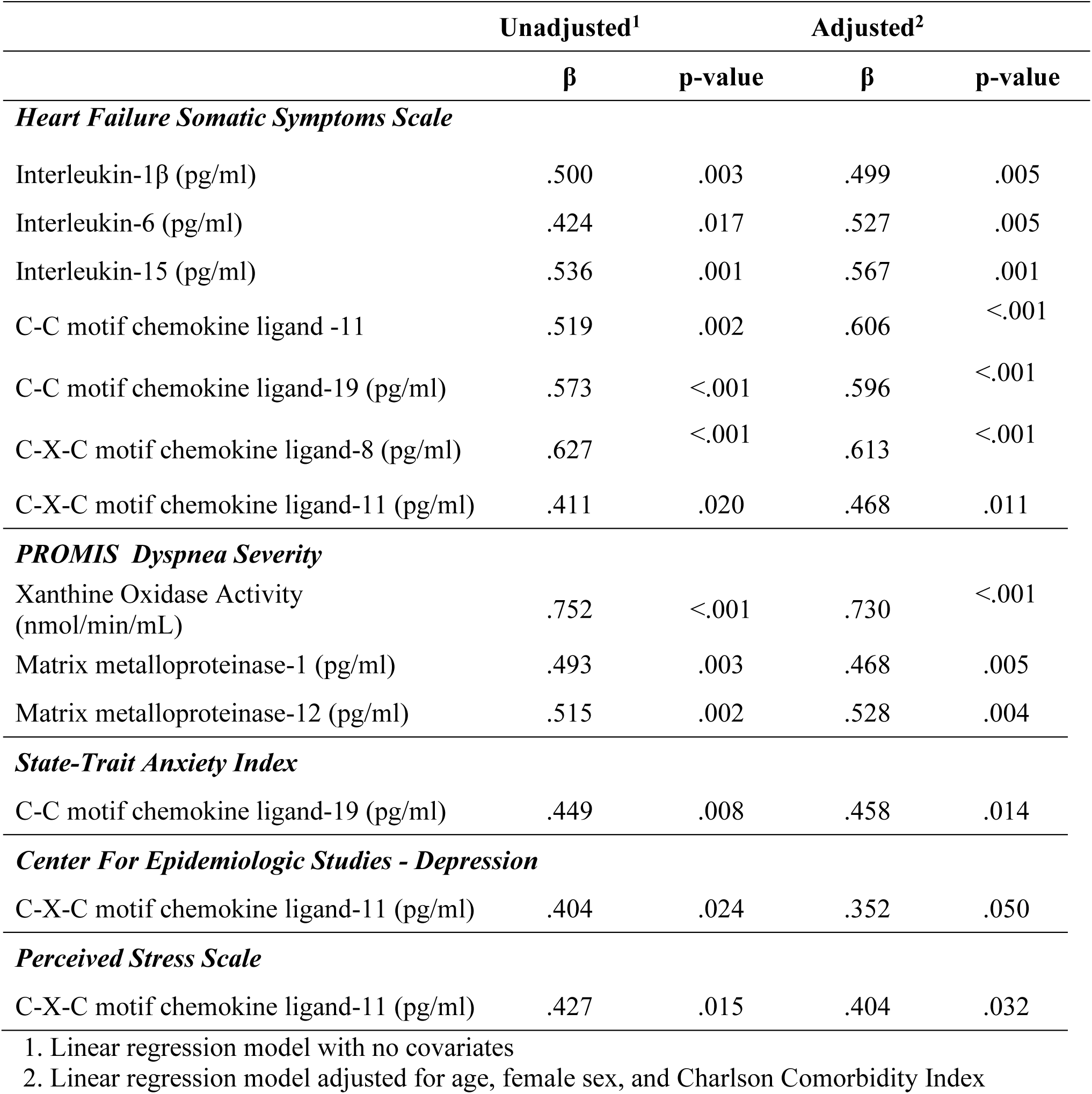
Adjusted and Unadjusted Effect Size Estimates for Biomarkers with Heart Failure and Psychological Symptoms.

HFSPS was positively related to cytokines associated with T-cell activation and differentiation, IL-1β, IL-6, and IL-15, and with chemokines involved in T cell recruitment, CCL-11, CCL-19, CXCL-8, and CXCL-11 (**Table 5**). The HFSPS Dyspnea, Subtle Early Symptoms, and Chest Discomfort subscales were positively related to T cell-related cytokines and chemokines (**Supplementary Table 2**).

## Discussion

This study found that biomarkers of oxidative stress, inflammation, vascular activation, fibrosis, and T cell activation were associated with physical symptoms. Chemokines associated with T cell activation were significantly related to psychological symptoms in Black adults with HF. To our knowledge, this is the first study to examine physiologic correlates of physical and psychological symptoms among Black adults. Physical symptoms in HF such as dyspnea, fatigue, and edema, are profoundly distressing due to their multifaceted impact on an individual’s quality of life and daily functioning. These symptoms directly correlate with HF progression, causing significant limitations in physical activity and leading to increased hospitalization and mortality rates. ^29^ Moreover, the chronic and often unpredictable nature of these symptoms exacerbates psychological distress, anxiety, and depression in individuals already coping with the burden of a chronic, life-threatening condition. ^30^ Understanding the intricate pathophysiologic pathways linked to symptoms in HF is crucial for developing targeted interventions and therapies, enabling precise management strategies that directly address the underlying mechanisms driving symptomatology.

This study found that elevated XO activity was related to higher dyspnea severity in Black persons with HF. Although further research in a larger sample is needed, these results could indicate new treatment options targeting chronic and oxidative stress for this population. Pharmacologic treatment targeting oxidative stress may include XO inhibitors, such as allopurinol and febuxostat, which have been consistently studied in the context of HF and shown to decrease uric acid levels and lower hospitalization rates.^21,22^ As dyspnea is a distressing symptom for persons living with HF, further work testing the effects of XO inhibitors on dyspnea severity and other HF symptoms is needed.

The association between dyspnea and elevated MMPs in individuals with HF underscores a potential mechanistic link between symptomatology and underlying cardiac remodeling processes. Understanding this relationship could identify the role of MMPs in pulmonary congestion and vascular changes, offering insights into novel therapeutic targets to alleviate symptoms and potentially modify the progression of HF.

The association between HF somatic symptoms (HFSPS) with inflammatory biomarkers and T cell activation markers signifies a potential link between immune system dysregulation and HF symptomatology. Understanding these relationships could link the role of inflammation in exacerbating symptoms, fostering new insights into targeted interventions aimed at modulating immune responses to alleviate distressing symptoms and potentially alter the trajectory of HF progression. ^31^

The association between symptoms of psychological distress (anxiety, depression, and perceived stress) and chemokines in Black individuals with HF suggests a potential link between mental health and immune signaling pathways, highlighting the intricate interplay between psychological factors and immune responses. Addressing psychological symptoms could potentially modulate immune processes, thereby influencing the symptom burden and disease progression in HF. ^32^ A greater understanding of these connections might offer insights into novel therapeutic approaches targeting both mental health and immune pathways to improve patient outcomes.

Higher neighborhood deprivation scores (ADI) were associated with higher perceived stress and higher PROMIS Sleep Deprivation scores among Black adults with HF, underscoring the significant impact of socioeconomic factors on psychological well-being within this population. This is further underscored by the association between higher ADI scores and higher inflammatory biomarkers in this population. These associations highlight the need for targeted interventions addressing social determinants of health to alleviate stress, ameliorate sleep disparities, reduce inflammation, thus potentially improving cardiovascular outcomes in this population.

## Limitations

This study was rolled out during the COVID-19 pandemic, leading to several limitations.

One, for the health and safety of study participants, this study was transitioned to a remote protocol collecting dried blood spots rather than whole blood, which excluded collection of PBMCs and genetic data. Thus, additional analyses related to immune cell function and potential genetic contributions could not be performed. Two, recruitment and enrollment efforts were delayed, and the ongoing pandemic was a barrier to meeting enrollment goals (N=50) upon implementation of the remote protocol. Three, this study was unable to account for the potential effects of the ongoing pandemic on inflammation and immune function. While we collected data on COVID-19 exposure and disease, with only 2 participants reporting prior COVID-19 infection, these data were self-reported. Replication studies with larger sample sizes will be needed to confirm the validity and generalizability of the results. Furthermore, while the study was able to find a positive relationship between inflammatory biomarkers and symptoms, more research is needed to uncover the underlying pathophysiologic mechanism for this association.

Notably, the findings of this study highlight the ‘hidden’ role that social determinants of health may play in the inflammatory process, thereby predisposing Black adults with HF to more physical and psychological symptoms. The American Heart Association advocates for the screening of social determinants of health as a routine part of clinical HF care. ^33^ In addition to the Area Deprivation Index, the Accountable Health Communities’ Social Needs Screening Tool is an easy-to-use, 10-item survey that assesses unmet needs pertaining to housing uncertainty, food insecurity, transportation availability, basic utilities, and interpersonal safety. ^34^ Standardized, routine clinical assessment of social determinants of health which impact the lived environment and are representative of access to care and socioeconomic status may provide an inclusive picture of the vulnerabilities which promote chronic stress, poor sleep, and systematic inflammation. Identification and subsequent mitigation of these risk factors during each clinical visit is vital to reducing the risk of chronic inflammation and its detrimental effect on HF symptoms.

## Conclusions

This study in Black adults with HF found associations between biomarkers of oxidative stress, inflammation, vascular activation, fibrosis, T cell activation and both physical and psychological symptoms. Physical symptoms, like dyspnea and edema, have a multifaceted impact on quality of life and are directly linked to HF progression, while psychological distress exacerbates symptoms and affects adherence to treatment. Elevated oxidative stress markers were linked to increased dyspnea severity, suggesting potential treatment avenues, while inflammatory biomarkers correlated with reduced quality of life, emphasizing the need to target inflammatory pathways. Furthermore, socioeconomic factors like neighborhood deprivation influenced stress, sleep, and inflammation in this population, emphasizing the importance of addressing social determinants of health.

## Data Availability

All data produced in the present study are available upon reasonable request to the authors.

## Acknowledgements

The study was supported in part by the National Institutes of Health grant numbers P30NR018090 and U54AG062334. Effort for this work was supported by National Institutes of Health grant numbers K23AG076977 (Butts) and T32NR012715 (Herring). The content is the sole responsibility of the authors and does not necessarily represent the official views of the National Institutes of Health.

## List of Abbreviations

ADI: Area Deprivation Index
CCL: C-C motif chemokine ligand
CES-D: Center for Epidemiological Studies Depression
CTRP1: C1q/tumor necrosis factor-related protein 1 (CTRP1
CXCL: C-X-C motif chemokine ligand
HF: heart failure
HFSPS: Heart Failure Somatic Perception
Scale IL: interleukin
LVEF: left ventricular ejection fraction
MFI: Multidimensional Fatigue Inventory
MMP: matrix metalloproteinase
NYHA: New York Heart Association
PROMIS: Patient-Reported Outcomes Measurement Information System
PSS: Perceived Stress Scale
STAI: State-Trait Anxiety Index
VCAM: vascular cell adhesion molecule
XO: xanthine oxidase

---

**Figure 1.**
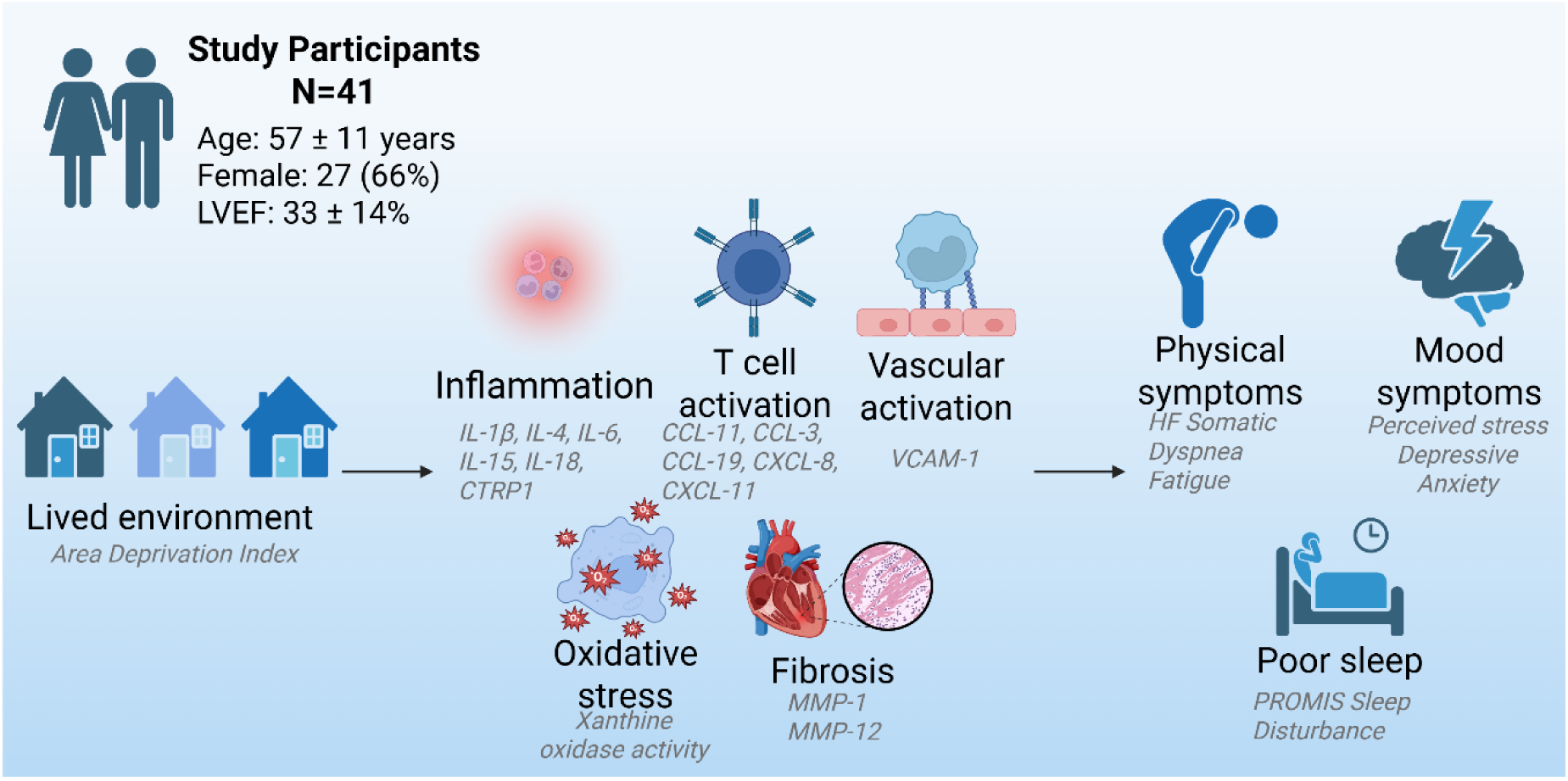
Relationships between inflammation, oxidative stress, and symptoms suggest potential therapeutic targets for alleviating heart failure burden. Addressing social determinants of health could reduce inflammation and improve cardiovascular outcomes in this population. Created in BioRender. Butts, B. (2024) https://BioRender.com/y71s620

**Supplementary Table 1.**
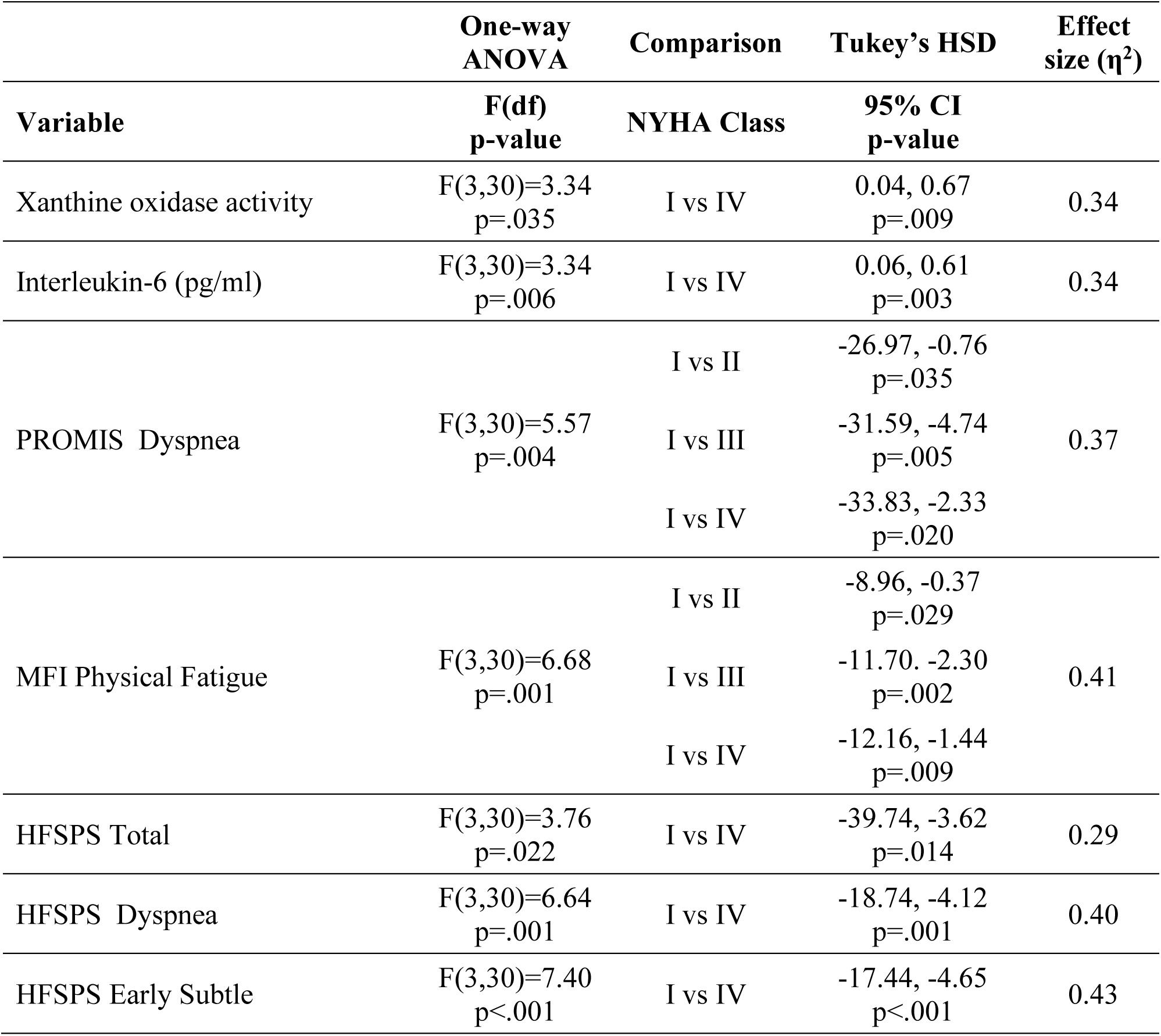
Biomarker Differences by NYHA Classification.

|  | **One-way**<br>**ANOVA** | **Comparison** | **Tukey's HSD** | **Effect**<br>**size ($\eta^2$)** |
| --- | --- | --- | --- | --- |
| **Variable** | **F(df)**<br>**p-value** | **NYHA Class** | **95% CI**<br>**p-value** |  |
| Xanthine oxidase activity | F(3,30)=3.34<br>p=.035 | I vs IV | 0.04, 0.67<br>p=.009 | 0.34 |
| Interleukin-6 (pg/ml) | F(3,30)=3.34<br>p=.006 | I vs IV | 0.06, 0.61<br>p=.003 | 0.34 |
| PROMIS Dyspnea | F(3,30)=5.57<br>p=.004 | I vs II | -26.97, -0.76<br>p=.035 | 0.37 |
|  |  | I vs III | -31.59, -4.74<br>p=.005 |  |
|  |  | I vs IV | -33.83, -2.33<br>p=.020 |  |
| MFI Physical Fatigue | F(3,30)=6.68<br>p=.001 | I vs II | -8.96, -0.37<br>p=.029 | 0.41 |
|  |  | I vs III | -11.70, -2.30<br>p=.002 |  |
|  |  | I vs IV | -12.16, -1.44<br>p=.009 |  |
| HFSPS Total | F(3,30)=3.76<br>p=.022 | I vs IV | -39.74, -3.62<br>p=.014 | 0.29 |
| HFSPS Dyspnea | F(3,30)=6.64<br>p=.001 | I vs IV | -18.74, -4.12<br>p=.001 | 0.40 |
| HFSPS Early Subtle | F(3,30)=7.40<br>p<.001 | I vs IV | -17.44, -4.65<br>p<.001 | 0.43 |

**Supplementary Table 2.** Heart Failure Perceived Somatic Symptoms Subscales Associations with Biomarkers.

| rho<br>p-value | Total | Dyspnea | Early | Chest | Edema | IL-1β | IL-6 | IL-15 | CCL-11 | CCL-19 | CXCL-8 | CXCL-11 |
| --- | --- | --- | --- | --- | --- | --- | --- | --- | --- | --- | --- | --- |
| **Total** | 1 | - | - | - | - | - | - | - | - | - | - | - |
| **Dyspnea** | .676<br>**<.001** | 1 | - | - | - | - | - | - | - | - | - | - |
| **Early** | .747<br>**<.001** | .718<br>**<.001** | 1 | - | - | - | - | - | - | - | - | - |
| **Chest** | .717<br>**<.001** | .693<br>**<.001** | .527<br>**<.001** | 1 | - | - | - | - | - | - | - | - |
| **Edema** | .361<br>**.036** | .534<br>**<.001** | .535<br>**<.001** | .385<br>**.020** | 1 | - | - | - | - | - | - | - |
| **IL-1β** | .500<br>**.020** | .444<br>**.009** | .498<br>**.003** | .017<br>.923 | .185<br>.294 | 1 | - | - | - | - | - | - |
| **IL-6** | .491<br>**.005** | .442<br>**.010** | .446<br>**.009** | .515<br>**.002** | .107<br>.552 | .138<br>.460 | 1 | - | - | - | - | - |
| **IL-15** | .536<br>**.001** | .414<br>**.012** | .325<br>.058 | .426<br>.101 | .100<br>.563 | .442<br>**.009** | .535<br>**<.001** | 1 | - | - | - | - |
| **CCL-11** | .519<br>**.002** | .419<br>**.012** | .324<br>.058 | .427<br>**.010** | .281<br>.102 | .332<br>.059 | .164<br>.340 | .292<br>.080 | 1 | - | - | - |
| **CCL-19** | .573<br>**<.001** | .331<br>**.052** | .511<br>**.002** | .312<br>.068 | .278<br>.106 | .230<br>.199 | .524<br>**.001** | .468<br>**.004** | .337<br>**.045** | 1 | - | - |
| **CXCL-8** | .627<br>**<.001** | .365<br>**.034** | .398<br>**.020** | .586<br>**<.001** | -.083<br>.062 | .267<br>.140 | .425<br>**.010** | .550<br>**<.001** | .309<br>.063 | .230<br>.177 | 1 | - |
| **CXCL-11** | .411<br>.020 | .430<br>.011 | .369<br>.032 | .371<br>.031 | .155<br>.382 | .150<br>.414 | .317<br>.056 | .135<br>.426 | .278<br>.096 | .168<br>.326 | .244<br>.146 | 1 |

